# A highly sensitive and specific SARS-CoV-2 spike- and nucleoprotein-based fluorescent multiplex immunoassay (FMIA) to measure IgG, IgA and IgM class antibodies

**DOI:** 10.1101/2021.07.28.21260990

**Authors:** Anna Solastie, Camilla Virta, Anu Haveri, Nina Ekström, Anu Kantele, Simo Miettinen, Johanna Lempainen, Pinja Jalkanen, Laura Kakkola, Timothée Dub, Ilkka Julkunen, Merit Melin

## Abstract

**Background:** Validation and standardization of accurate serological assays are crucial for the surveillance of the coronavirus disease 2019 (COVID-19) pandemic and population immunity.

**Methods:** We describe the analytical and clinical performance of an in-house fluorescent multiplex immunoassay (FMIA) for simultaneous quantification of antibodies against the severe acute respiratory syndrome coronavirus 2 (SARS-CoV-2) nucleoprotein and spike glycoprotein. Furthermore, we calibrated IgG-FMIA against World Health Organisation (WHO) International Standard and compared FMIA results to an in-house enzyme immunoassay (EIA) and a microneutralisation test (MNT). We also compared the MNT results of two laboratories.

**Results:** IgG-FMIA displayed 100% specificity and sensitivity for samples collected 13-150 days post-onset of symptoms (DPO). For IgA- and IgM-FMIA 100% specificity and sensitivity were obtained for a shorter time window (13-36 and 13-28 DPO for IgA- and IgM-FMIA, respectively). FMIA and EIA results displayed moderate to strong correlation, but FMIA was overall more specific and sensitive. IgG-FMIA identified 100% of samples with neutralising antibodies (NAbs). Anti-spike IgG concentrations correlated strongly (ρ=0.77-0.84, *P*<2.2×10^−16^) with NAb titers. The NAb titers of the two laboratories displayed a very strong correlation (ρ=0.95, *P*<2.2×10^−16^).

**Discussion:** Our results indicate good correlation and concordance of antibody concentrations measured with different types of in-house SARS-CoV-2 antibody assays. Calibration against WHO international standard did not, however, improve the comparability of FMIA and EIA results.

## Introduction

Severe acute respiratory syndrome coronavirus 2 (SARS-CoV-2), the causative agent of coronavirus disease 2019 (COVID-19), had claimed over 4 million lives and infected 190 million by late July 2021 [1]. A large proportion of all infections may go undetected in their acute phase [2,3] for various reasons, such as lack of symptoms [4,5] or hesitancy of getting tested [6]. Therefore, accurate serological assays are needed to provide more reliable estimates of COVID-19 prevalence.

SARS-CoV-2 serological assays are useful anywhere between determining seroprevalence in the general population to the investigation of confined outbreaks. In neutralisation tests and other serological tests alike, the clinical specificity and sensitivity of the assays can vary considerably [7–12]. Comparisons of methods and their standardisation is urgently needed to properly understand and apply the vast acquired information on the immunity induced by SARS-CoV-2 infection and COVID-19 vaccines.

We describe the validation and performance of an in-house fluorescent multiplex immunoassay (FMIA) developed for quantification of antibodies produced against three SARS-CoV-2 antigens: the full-length spike glycoprotein (SFL), spike receptor-binding domain (RBD) and nucleoprotein (N). Our assay detects antibodies against these three antigens simultaneously, which enables differentiation of natural SARS-CoV-2 infection from vaccine-induced immunity. The assay is based on an FMIA previously described by Trivedi et al. [13]. We have recently reported the performance of the FMIA for measuring IgG antibodies against SARS-CoV-2 nucleoprotein [14]. Here, we report our findings on the analytical and clinical performance of FMIA and compare its IgG, IgA and IgM assay results to another laboratory’s in-house enzyme immunoassay (EIA) [15,16], both calibrated for IgG with WHO International Standard [17]. Furthermore, we compare FMIA antibody levels to neutralizing antibody (NAb) titers [14,18] and NAb titers of microneutralisation tests (MNTs) between two separate laboratories.

## Materials and methods

### Serum samples

We assessed the analytical and clinical performance of the FMIA by using negative and positive sample panels. The negative sample panel consisted of 402 anonymous serum samples collected in mid-2019 before the COVID-19 pandemic. The positive serum panel consisted of 147 samples collected from 58 volunteers 4–150 days post-onset of symptoms (DPO) and 0–147 days post positive PCR test result for SARS-CoV-2. Samples were collected as part of a COVID-19 household transmission study [19]. We used the negative and positive serum panels also to compare FMIA and MNT results. We excluded one sample due to limited sample volume leaving the total sample size to n=548. Details of the panels are presented in Table S1 and Figure S1.

We compared FMIA with EIA using a serum panel that consisted of a subset of 80 samples collected as part of a COVID-19 mRNA vaccine response study [16]. We collected the samples from convalescent-phase PCR-confirmed COVID-19 patients (n=20) and volunteer health care workers (n=20) who received two doses of Pfizer-BioNTech BNT162b2 mRNA vaccine. We recruited the convalescent COVID-19 patients among outpatients with a positive RT-PCR positive test result in the Helsinki University Hospital database and collected serum samples 15-41 (median 26) DPO. We collected sera from 20 vaccinated volunteers before or on the day of the vaccination (day 0 samples), three weeks (median 21 days, range 18-21) post 1^st^ vaccine dose (3-week samples), and six weeks (median 48 days, range 39-50) post 1^st^ vaccine dose (6-week samples). At the time of the 6-week sample collection, three weeks (median 27 days, range 18-29) had also passed from the 2^nd^ vaccine dose. Participant demographics are presented in Table S1.

### The SARS-CoV-2 FMIA

Here, we adapted the previously described SARS-CoV-2 FMIA [14] for the measurement of IgG, IgA and IgM antibodies against SARS-CoV-2 RBD, SFL and N antigens. Briefly, we conjugated SARS-CoV-2 RBD (product code: REC31849), SFL (REC31868) and N (REC31812, the Native Antigen Company) antigens on the surfaces of MagPlex®-C superparamagnetic carboxylated microspheres (Luminex) by carbodiimide reaction. We added the microspheres onto black 96-well plates (Costar 3915) with diluted sera, reference and control samples and incubated RT with shaking at 600 rpm in the dark for 1 hour. We washed the unbound particles away with a magnetic plate washer (ELx405 and 405TSRS, BioTek), and added R-Phycoerythrin-conjugated Affinipure Goat Anti-Human IgG, IgA or IgM Fcγ Fragment Specific detection antibodies (Jackson Immuno Research) to the wells. We incubated the plates for 30 minutes as described above and then washed them again. We measured the median fluorescence intensity (MFI) with MAGPIX® system (Luminex). MFI values were automatically converted into antibody concentration (FMIA U/ml) via interpolation from 5-parameter logistic (5-PL) curves (xPONENT software version 4.2, Luminex) created from serially diluted (1:400–1:1638400) in-house reference serum. We gave the 1:400 dilution of the standard an arbitrary concentration of 100 FMIA U/ml. We performed the FMIA analyses at the Finnish Institute for Health and Welfare (Helsinki, Finland). A full description of the FMIA method is presented in Supplementary Material.

### Evaluation of the analytical performance of FMIA

#### Limit of detection (LOD) and limit of quantification (LOQ)

We determined LOD and LOQ from 26 experiments by calculating the mean + 3x (LOD) and 8x (LOQ) standard deviation of MFI values generated from blank wells (n=52). MFI values were converted into FMIA U/ml by interpolation from the 5-PL reference curve with GraphPad Prism v9.

#### Linearity

We assessed the linearity of FMIA by comparing antibody concentration of serially diluted (1:100, 1:200, 1:400, 1:800 and 1:1600) sera (IgG: n=6, IgA and IgM: n=7) collected as part of the COVID-19 household transmission study [19]. Linearity was measured as Pearson correlation between dilution factor and antibody concentration, and mean R^2^≥0,95 of all samples was considered acceptable.

#### Precision and reproducibility

We assessed the precision and reproducibility of FMIA as the mean percentage of the coefficient of variation (CV%) of antibody concentrations between samples analysed repeatedly in different settings. Precision was assessed as intra- and inter-assay variation and reproducibility by comparing the results obtained by three laboratory technicians and as variation caused by different batches of crucial reagents. A full description of the evaluation of analytical performance is presented in Supplementary Material.

#### Evaluation of the clinical performance of FMIA

We determined the clinical performance of the antibody assays as their ability to distinguish negative- (pre-COVID19 era) and positive panels’ (PCR confirmed patients’) sera. We determined the thresholds for positivity by comparing all possible threshold combinations for the three antigens with R (version 3.6.0) and RStudio (version 1.2.1335). We prioritised a specificity of 100% and determined it with the entire negative serum panel (n=402). Sensitivity was assessed separately for subgroups of the positive serum panel’s samples (n=147) based on DPOs.

#### EIA

The SARS-CoV-2 EIA used in this study has previously been described in detail [15]. We measured IgG, IgA, IgM and total Ig antibodies against SARS-CoV-2 S1 and N proteins from sera diluted 1:300. SARS-CoV-2 S1 (GenBank ID: MN908947.3) and N (Genbank ID: NC_045512.2) antigens were expressed and purified, and antigen-specific antibody levels were measured with a Victor Nivo device (PerkinElmer) as described previously [16]. We converted the results into EIA units by comparing the absorbance values of samples with the absorbance values of 1:300 diluted positive control (marked as 100 EIA units) and 1:300 diluted negative control (marked as 0 EIA units) included in each assay. Thresholds for positivity were previously determined with 20 negative sera [16]. We performed the EIA analyses at the University of Turku (Turku, Finland).

#### Calibration against WHO International Standard

We calibrated the FMIA’s IgG specific in-house reference serum against a WHO International Standard (NIBSC code 20/136 [17]). We serially diluted the in-house reference serum and WHO international standard and assigned the 1:100 dilution of the WHO international standard a concentration of 10 binding antibody units (BAU)/ml. We conducted the analysis on two separate days and interpolated the mean concentration for each dilution of the in-house reference serum from the linear range of the WHO international standard. With EIA, we used the WHO international standard to calibrate the positive control by diluting both 1:300 and calculating their ratio. We performed the EIA analysis once and used the average absorbance values calculated from duplicate wells. We obtained a calibration factor for each antigen separately and used them to convert FMIA U/ml and EIA units into BAU/ml.

#### Microneutralisation tests

##### MNT of laboratory #1

The MNT of laboratory #1 was performed at the Finnish Institute for Health and Welfare (Helsinki, Finland) and the assay has been previously described in detail [14,18]. Briefly, the MNT of laboratory #1 was performed using Vero E6 cells and true duplicates of sera diluted serially from 1:4. After four days the cells were fixed with 30% formaldehyde and the cytopathic effect (CPE) was measured. The viral strains used were hCoV-19/Finland/1/2020 (FIN-1) (GISAID accession ID EPI_ISL_407079) and hCoV-19/Finland/FIN-25/2020 (FIN-25) in the analysis of positive and negative serum panels, and FIN-1 in analyses of sera used in comparison to MNT of laboratory #2. A sample was considered positive when the NAb titer was ≥6 for at least one virus and negative when titers for both viruses were <4. Samples with titers between 6 and 4 for both viruses were considered borderline. Negative samples were given a titer value of 2 for statistical analyses. In cases where the titers of the two viruses differed, the titer of the sample was defined as the highest of the two.

##### MNT of laboratory #2

The MNT of laboratory #2 was performed at the University of Turku (Turku, Finland) and the method has been previously described [16]. The MNTs between laboratories 1 and 2 differed in used viral isolate, serum dilution, cell line, incubation time and cell fixation. MNT of laboratory #2 used VeroE6-TMPRSS2-H10 cells fixed with 4% formaldehyde and measured CPE after three days. Starting dilution for sera was 1:20 with 50 TCID_50_ of SARS-CoV-2 variant FIN-25 (EPI_ISL_412971, GenBank accession MW717675) and neutralisation titers were analysed in duplicates. The NAb titer was defined as the last dilution resulting in 50% inhibition of cell death. The threshold for positivity was ≥20. Negative samples were given a titer value of 10 for statistical analyses. FIN-1 represents lineage B (D614G substitution) and FIN-25 represent lineage B.1 with three amino acid substitutions (D614G, 41% R682W, 45% YQTQT 674-678 [16]) in the spike glycoprotein compared to the Wuhan-Hu-1 strain.

##### Comparison of FMIA, EIA and MNT assays

We calculated Spearman correlation coefficient (ρ) and the statistical significance of the correlation between FMIA and EIA antibody concentrations and the two laboratories’ NAb titers. In addition, we compared FMIA and EIA in their ability to detect antibodies induced by natural infection and vaccination. Samples that were borderline in MNT of laboratory #1 were considered as positives.

##### Ethical statement

The investigations were carried out under the General Data Protection Regulation (Regulation (EU) 2016/679, directive 95/46/EC) and the Finnish Personal Data Act (Finlex 523/1999). The study protocols for the collection of pre-COVID-19 pandemic samples and COVID-19 patient samples were approved by the Helsinki-Uusimaa health district ethical permissions 433/13/03/00/15 and HUS/1238/2020. The Finnish law on communicable diseases and the duties of the Finnish Institute for Health and Welfare [20,21] allowed the implementation of the initial household transmission study without seeking further institutional ethical review. A waiver of ethical approval was received from Prof. Mika Salminen (Director of the Department for Health Security of the Finnish Institute for Health and Welfare). The vaccinee cohort (n=20) was randomly selected from a larger cohort (n=180) of vaccinated healthcare personnel of Turku University Hospital [16] approved by the Southwest Finland health district ethical permission ETMK 19/1801/2020. Written informed consent was obtained from all volunteers.

## Results

### Analytical performance of the FMIA

We calculated the LOQ and LOD separately for each antigen and each antibody class and the data is presented in Table S2. For IgA and IgM assays, the linearity with different serum dilutions was excellent for all antigens (range R^2^=0.96-1). For the IgG assay, less diluted samples (dilutions 1:100 and 1:200) resulted in relatively lower IgG concentrations leading to weaker linearity correlations. Exclusion of dilutions 1:100 and 1:200 resulted in R^2^≥0.99 for all antigens in the IgG assay. As a compromise to avoid decreased clinical sensitivity and to minimize the number of serum dilutions, we decided to calculate antibody concentrations from the average of 1:100 and 1:1600 dilutions for all except negative sample panels sera, which analysed diluted 1:100.

The mean intra-assay variation ranged from 8 to 10% and inter-assay variation from 4 to 12% in the different antibody classes (Table S3). The mean variation between different technicians was 20% for IgG, 21% for IgA and 18% for IgM assays. The variation between four different batches of conjugated microspheres was 16% and the variation between different batches of detection antibodies was 15% for IgA and IgG assays. Overall, both intra- and inter-assay variation were found acceptable for all antibody classes and antigens.

### Calibration against WHO international standard

The IgG specific concentrations against N, RBD and SFL of the in-house reference serum calibrated against the WHO International standard [17] were 34, 18 and 23 BAU/ml, respectively. Because the starting dilution of IgG specific in-house reference sera was given an arbitrary concentration of 100 FMIA U/ml we multiplied the results by a factor of 0.34 for nucleoprotein, 0.18 for RBD and 0.23 for SFL to obtain calibrated BAU/ml. In EIA, the calibration factor for N IgG was 12.9, for N total Ig 17.4, for S1 IgG 11.8 and for S1 total Ig assay 12.0. We did not calibrate MNTs but obtained a titer of 192 against FIN-1 virus in laboratory #1 and 640 against FIN-25 in laboratory #2 for the WHO international standard.

### Clinical performance of the FMIA

We set the thresholds for positivity based on a positive panel of convalescent sera (n=147) and a negative serum panel (n=402) aiming to achieve 100% specificity. We considered a sample positive for anti-spike IgG antibodies (S-IgG) if it had ≥0.13 and ≥0.089 BAU/ml anti-RBD and anti-SFL IgG antibodies, respectively. With these thresholds, the specificity of the S-IgG FMIA was 100% [95% CI: 99.1–100] (Table 1). The sensitivity of the FMIA for detecting antibodies in convalescent sera was dependent on DPO (Table 1). The sensitivity of the S-IgG FMIA assessed based on the positive serum panel (DPO 4– 150, n=147) was 97% [95% CI: 93.2–98.9]. All samples with IgG concentrations under the thresholds (n=7/147) were collected 4–11 DPO (Figure 1). The sensitivity of the S-IgG FMIA was 100% [95% CI: 97.3–100] for samples collected 13–150 DPO (n=140) (Table 1). With the positive serum panel used in this study (n=147) the specificity and sensitivity of the N-IgG FMIA we have previously described [14] was 100% for samples collected 21-51 DPO but the sensitivity of the assay decreased to 98% for samples collected 52-150 DPO (Figure 2).

**Table 1.**
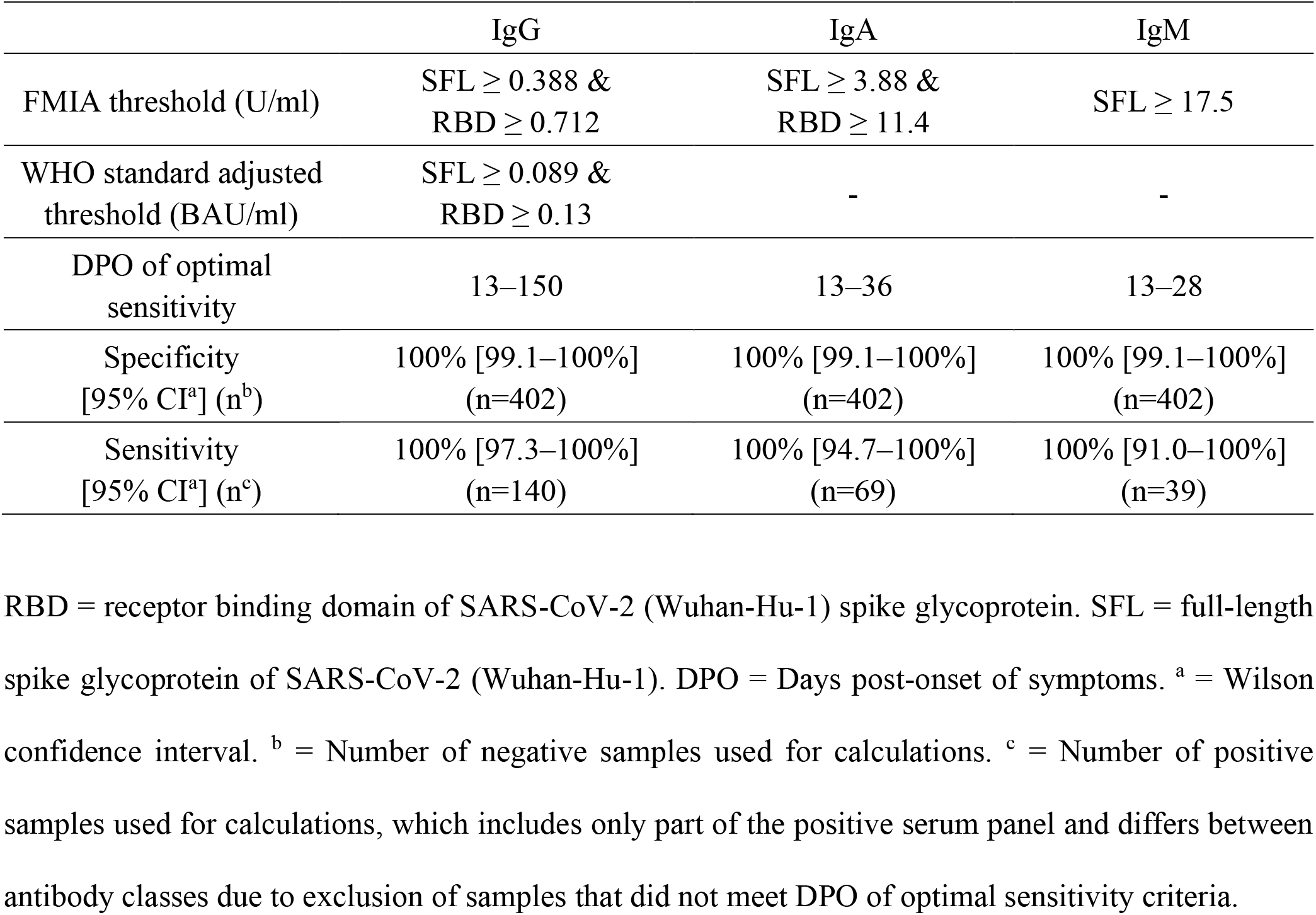
Thresholds, clinical specificity and clinical sensitivity of FMIA.

**Figure 1.**
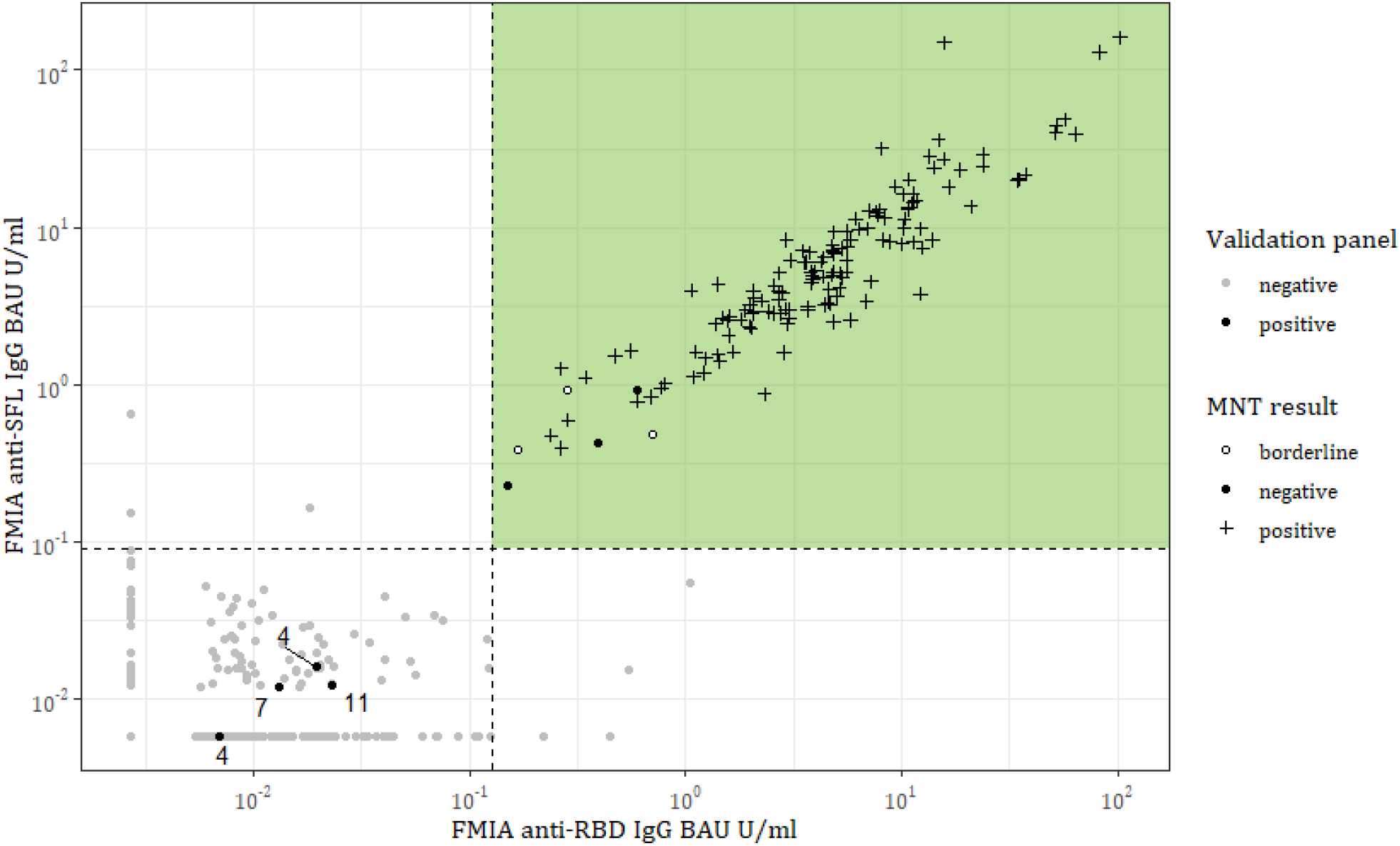
Anti-SFL and RBD IgG antibody concentrations (BAU/ml) of positive (black, n=147) and negative (grey, n=402) serum panels measured with FMIA. Dashed lines represent IgG SFL and RBD assay thresholds, and samples that pass both thresholds (coloured area) are classified as positive for SARS-CoV-2 spike specific IgG antibodies. The numbers indicate days post-onset of symptoms for samples that belong to the positive serum panel but were categorized as negative for anti-spike IgG antibodies in FMIA. MNT result = microneutralisation test interpretation; <4 = negative, 4 = borderline, >4 = positive. RBD = receptor binding domain of SARS-CoV-2 (Wuhan-Hu-1) spike glycoprotein. SFL = full-length spike glycoprotein of SARS-CoV-2 (Wuhan-Hu-1).

**Figure 2:**
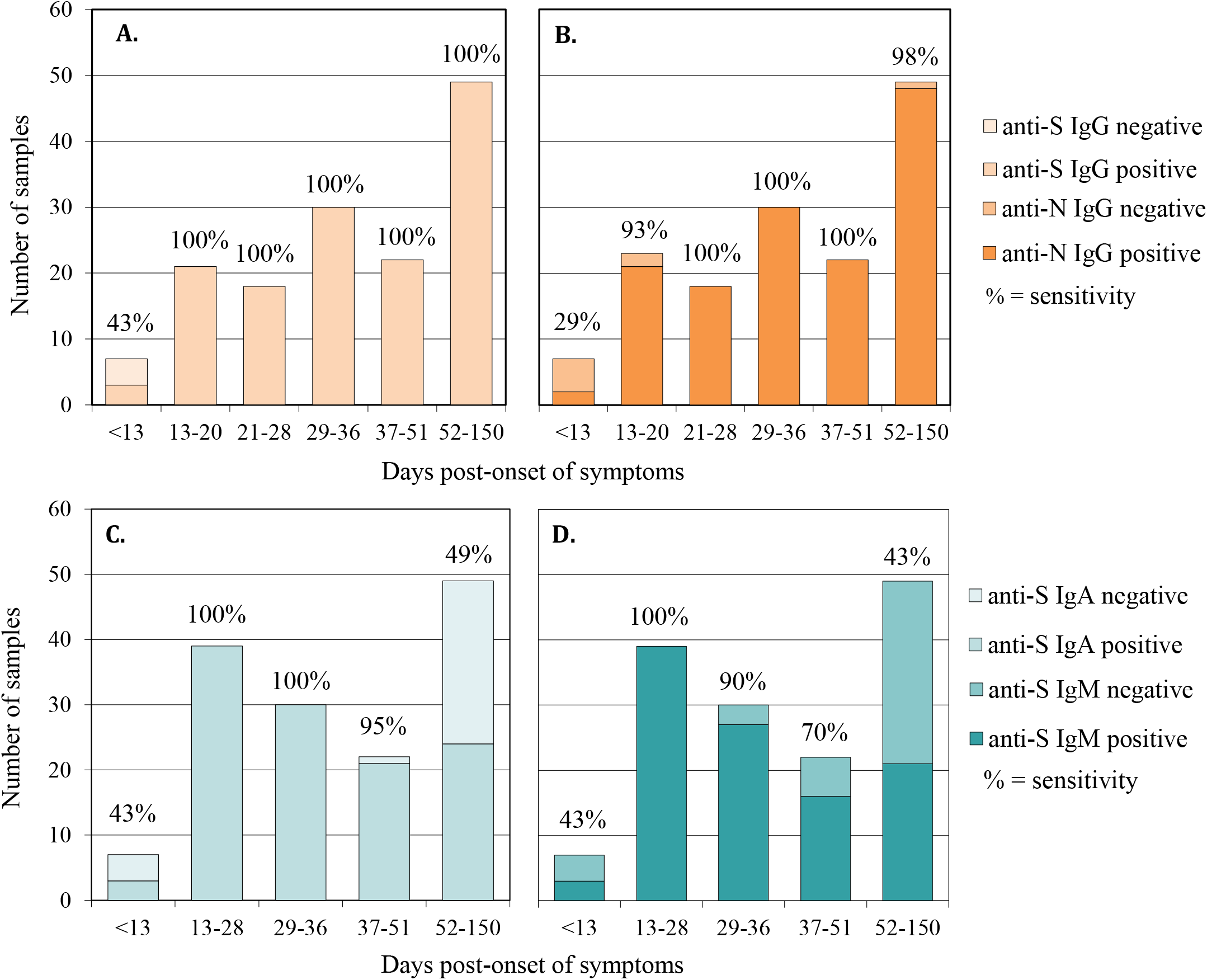
Sensitivity (%) of SARS-CoV-2 FMIA and the effect of time post-onset of symptoms (DPO). **A**. Anti-S (SFL and RBD) IgG FMIA. **B**. Anti-N IgG FMIA. **C**. Anti-S (SFL and RBD) IgA FMIA. **D**. Anti-S (SFL) IgM FMIA. n = number of positive serum panel’s samples in each DPO group. RBD = receptor binding domain of SARS-CoV-2 (Wuhan-Hu-1) spike glycoprotein. SFL = full-length spike glycoprotein of SARS-CoV-2 (Wuhan-Hu-1). N = SARS-CoV-2 nucleoprotein of SARS-CoV-2 (Wuhan-Hu-1).

The optimal threshold for IgA-FMIA was also based on the simultaneous detection of both spike antibodies. We considered a sample positive for IgA antibodies if it contained ≥11.4 and ≥3.88 FMIA U/ml anti-RBD and anti-SFL IgA antibodies, respectively, which resulted in 100% specificity [95% CI: 99.1–100] (Table 1.). For samples collected 13–36 DPO (n=69), the sensitivity was 100% [95% CI: 94.7–100]. As DPO increased, sensitivity decreased as a lower number of the positive serum panel’s samples reached the thresholds (Figure 2).

The optimal threshold for IgM-FMIA was based only on the concentration of anti-SFL antibodies, as the addition of RBD or N to the threshold criteria did not result in higher specificity or sensitivity. We considered samples that contained ≥17.5 FMIA U/ml anti-SFL IgM antibodies positive for IgM. The specificity of the IgM assay was 100% [95% CI: 99.1–100] with this threshold (Table 1). DPO range resulting in 100% sensitivity was 13–28 days for IgM [95% CI for sensitivity: 91.0–100, n=39] (Table 1). As DPO increased, the sensitivity of the IgM assay decreased steeply (Figure 2).

### Comparison of FMIA and MNT

NAb titers exhibited strong (ρ=0.77-0.84) and statistically significant correlation (*P*<2.2×10^−16^) with S-IgG concentrations (Figure 3). All samples that contained NAb were also positive in IgG-FMIA regardless of DPO (Table S4). Thereby the S-IgG FMIA was 100% [95% CI: 97.3-100] sensitive and 96% [95% CI: 91.1-98.4] specific for identification of NAb from samples taken 4-150 DPO. Some samples were negative in MNT but positive for S-IgG in FMIA (Table S4) indicating that not all antibodies that bind to SFL and RBD antigens used in FMIA are neutralising. The ability of IgA-and IgM-FMIA to identify samples positive for NAbs was dependent on DPO in a manner similar to their clinical sensitivity, indicating the different kinetics of IgA and IgM compared to NAb antibodies (Table S4, Figure 2).

**Figure 3.**
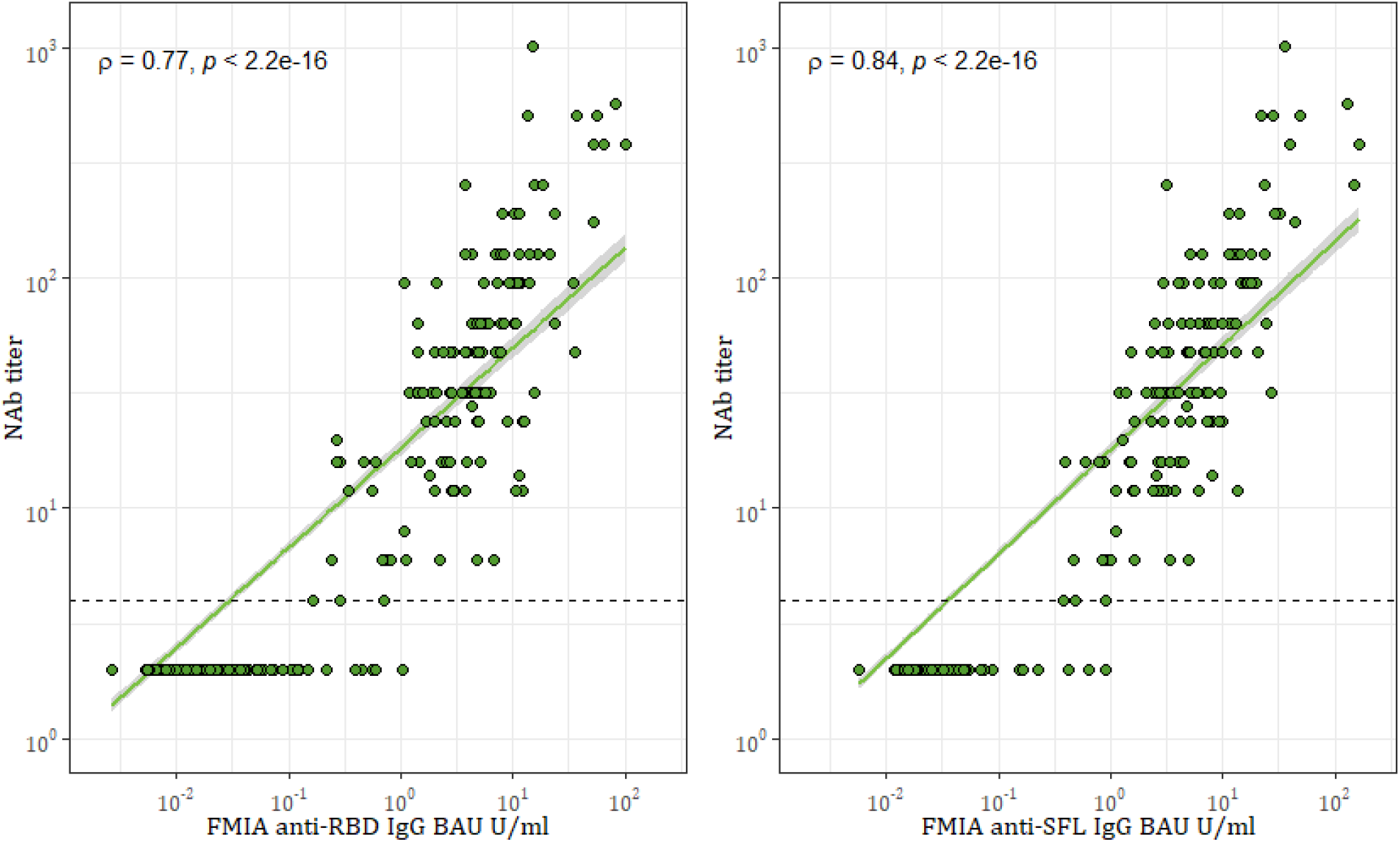
Spearman correlation (ρ) and significance (*p*) between FMIA anti-spike antibody concentrations and neutralising antibody (NAb) titers. The dashed line marks the threshold for positive MNT result (>4). One point may represent multiple samples (n=548). RBD = receptor binding domain of SARS-CoV-2 (Wuhan-Hu-1) spike glycoprotein. SFL = full-length spike glycoprotein of SARS-CoV-2 (Wuhan-Hu-1).

### Comparison of FMIA and EIA

We compared the results of FMIA and EIA by analysing a panel of convalescent serum samples (n=20) and pre-and post-vaccination samples (n=60) of 20 subjects. We observed the strongest correlations (ρ=0.94-0.95) between FMIA and EIA for IgG (FMIA) and IgG or total Ig (EIA) antibody concentrations against SARS-CoV-2 spike antigens (Figure 4). FMIA and EIA anti-spike antibody concentrations also displayed a strong correlation in IgA and IgM assays (ρ=0.79 for IgA, ρ=0.88 for IgM, Figure S2). The correlation between anti-N concentrations of IgG FMIA and EIA was moderate (ρ=0.54, Figure 4). Thus, FMIA and EIA differed to some extent in their ability to measure antibodies induced by SARS-CoV-2 infection and COVID-19 mRNA vaccination.

**Figure 4.**
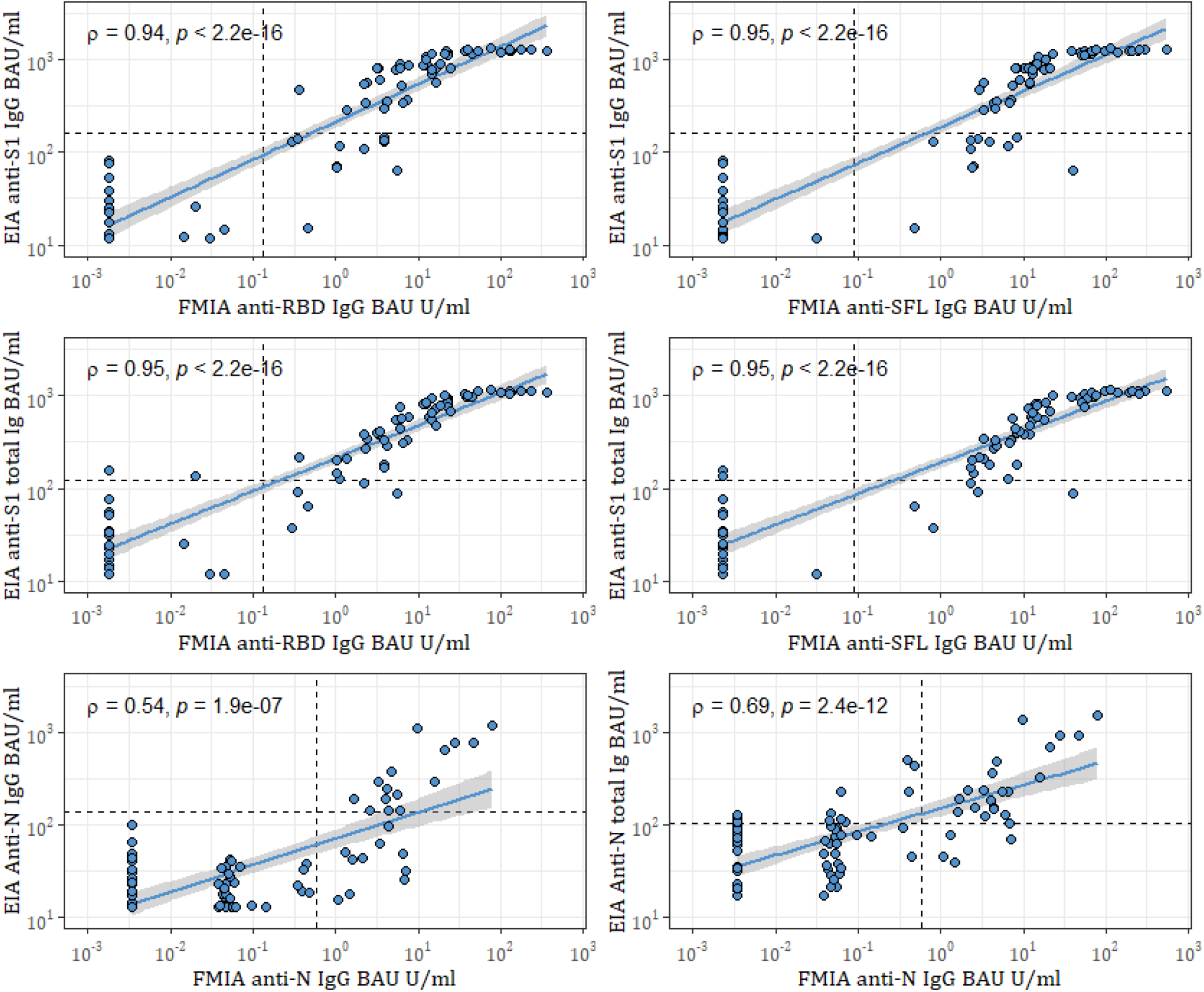
Spearman correlation (ρ) and significance (*p*) between FMIA IgG and EIA IgG and total anti-spike Ig antibodies in binding antibody units (BAU/ml) calibrated against WHO international standard. Dashed lines mark thresholds for positivity per antigen. One point may represent multiple samples (n=80). S1 = SARS-CoV-2 spike glycoprotein S1 subunit, RBD = receptor binding domain of SARS-CoV-2 (Wuhan-Hu-1) spike glycoprotein. SFL = full-length spike glycoprotein of SARS-CoV-2 (Wuhan-Hu-1). N = SARS-CoV-2 nucleoprotein of SARS-CoV-2 (Wuhan-Hu-1).

Of convalescent patient sera (14-60 DPO), 20/20 had anti-N and S-IgG antibodies in FMIA, while 15/20 had anti-N and 11/20 anti-S1 in IgG EIA and 19/20 had anti-N and 17/20 had anti-S1 antibodies in EIA total Ig assay. Anti-spike IgA antibodies were identified in 18/20 of patients when measured with FMIA but only 1/20 had anti-S1 IgA antibodies with EIA. Anti-spike IgM antibodies were identified in 18/20 and 20/20 of patients with FMIA and EIA, respectively. Of the samples collected before vaccination, 1/20 (IgG FMIA), 0/20 (IgG EIA) and 5/20 (EIA total Ig) had anti-N antibodies while lacking anti-spike antibodies. Of samples collected at 3 weeks after vaccination, 20/20 were positive for anti-spike IgG in FMIA and 18/20 in EIA; 20/20 samples were positive at 6 weeks with both assays.

When we compared the IgG concentrations in convalescent and post-vaccination samples measured with FMIA and EIA, the mean CV between FMIA U/ml and EIA units was 80%. The mean CV increased to 130% when results were converted to BAU/ml. Hence, calibration with the WHO standard did not increase the comparability of FMIA and EIA but increased the variation between the two assays instead.

### Comparison of MNTs

We compared the results of the NAb titers of two different laboratories with the same convalescent, pre- and post-vaccination serum panels as with FMIA and EIA. The correlation between NAb titers of the two laboratories was very strong at ρ=0.95 (*P*<2.2×10-16) (Figure 5). Both MNTs classified 20/20 day 0 samples as negative and 20/20 6-week post-vaccination samples as positive. The MNT of laboratory #1 was more sensitive at 3 weeks by identifying 18/20 of samples as having NAbs compared to 12/20 of laboratory #2. Among convalescent-phase patient sera, NAbs were found in 19/20 of samples in laboratory #1 and 18/20 of the samples in laboratory #2.

**Figure 5.**
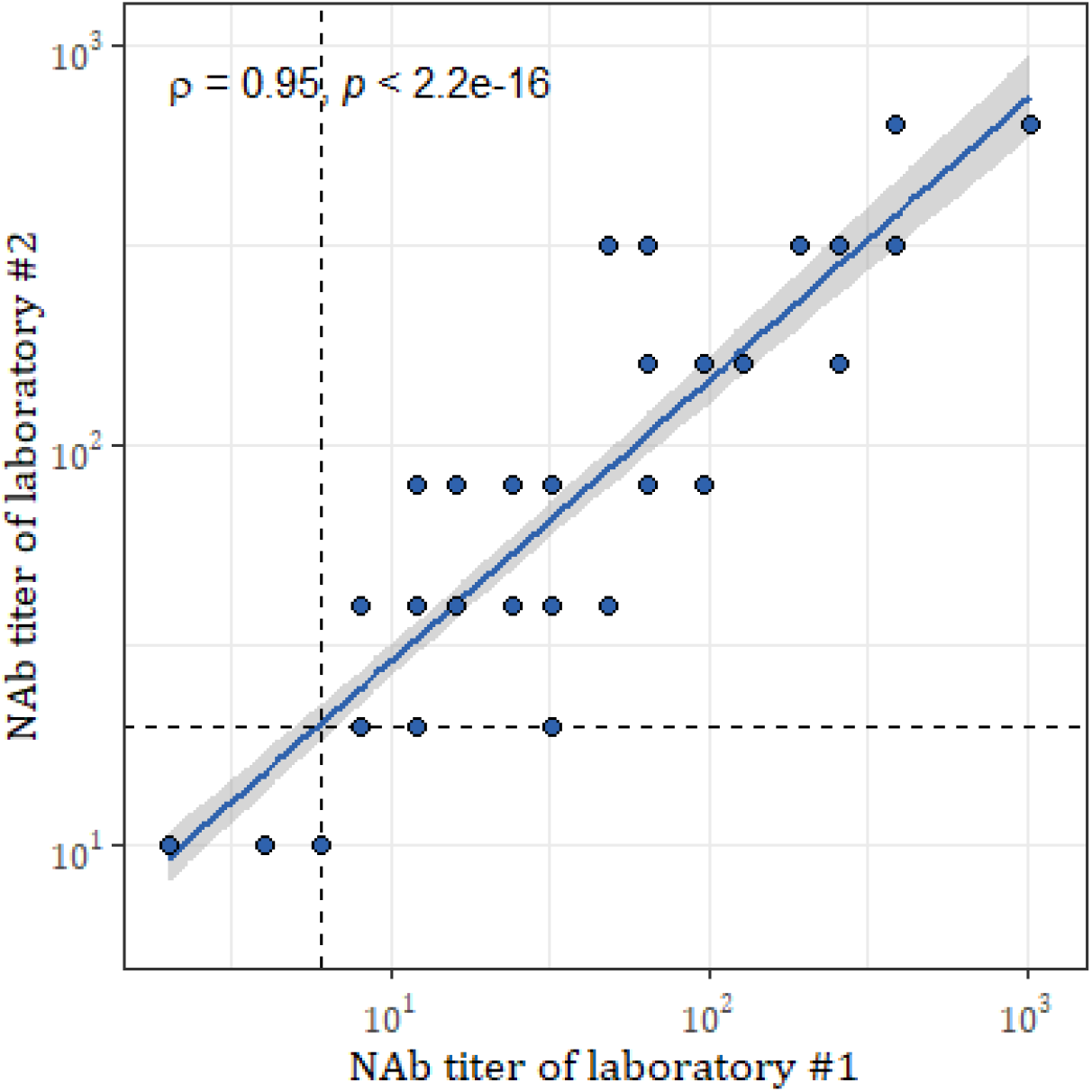
Spearman correlation (ρ) and significance (*p*) between neutralising antibody (NAb) titers of two laboratories. Dashed lines mark thresholds for positivity (≥6 for laboratory #1 and ≥20 for laboratory #2). One point may represent multiple samples (n=80).

## Discussion

We evaluated the analytical and clinical performance of SARS-CoV-2 FMIA and compared FMIA to other serological assays. The clinical performance of the FMIA was excellent, reaching 100% sensitivity and specificity. The clinical performance of S-IgG FMIA did not decrease through the sample collection period (150 days or 5 months). The sensitivity of IgA- and IgM-FMIA decreased with increasing DPOs, consistent with the shorter half-lives of IgA and IgM antibodies [22].

FMIA performed well in comparison to the in-house EIA antibody test. The correlation between FMIA and EIA IgG and total Ig results was strong for spike glycoprotein and moderate for nucleoprotein. Our results indicate that both methods accurately recognise vaccine-induced antibody responses at 6 weeks, but that FMIA is slightly more sensitive than EIA in samples taken 3 weeks after 1^st^ vaccine dose. As we compared the MNTs of two laboratories, we found that NAb titers had a very strong correlation despite some differences in the methodology. Overall, the MNT of one of the laboratories was somewhat more sensitive, which is likely explained by less diluted sera and different viral strains (lineage B vs. B.1).

Limitations of this study are related to the sample material used in the validation process of FMIA. In essence, the thresholds and DPOs described here were optimised for the detection of previous infections with mild to moderate symptoms. Despite this, the S-IgG levels correlated strongly with NAb titers. Our results also suggest that FMIA is better at recognising samples with NAbs than many commercial assays [12]. Whilst S-IgG FMIA identified all samples with NAbs, it does not necessarily measure antibodies against neutralising epitopes only, as some samples negative in MNT were considered positive for S-IgG. We conclude that FMIA is valuable in pre-screening of serum samples prior to confirmatory MNT. As the anti-SARS-CoV-2 antibody levels have been found to correlate with the disease severity [23–25], we can expect FMIA to perform well in the serological diagnostics of also previous severe infections. Importantly, while the present data show an excellent sensitivity for IgG-FMIA until 150 DPO, its performance after that remains to be investigated.

Estimates of the persistence of immunity to COVID-19 appear to depend on the serological assay used [11,26]. We recently reported that six and twelve months after SARS-CoV-2 infection 98% and 97 % of the patients, respectively, still had S-IgG in FMIA [27]. Other recent studies have found that anti-spike IgG antibodies persist in 90% of individuals seven [28] and nine months [29] after a confirmed SARS-CoV-2 infection. We found no decrease in the sensitivity of the spike-based IgG-FMIA in samples collected up to 5 months after infection. Anti-nucleoprotein antibodies have been reported to wane faster [27,30], consistent with our findings of sensitivity decreasing from 100 to 98% for N-IgG FMIA 52-150 DPO. Together with data of persisting antibodies, our findings imply, that S-IgG FMIA continues to be reliable in the detection of antibodies produced against SARS-CoV-2 for at least several months after infection.

Notably, the participants providing convalescent serum samples for the positive serum panel were likely all infected with a B.1 lineage virus since the samples were collected in Finland between March and September 2020. However, by using two spike antigens (RBD and SFL), both of which include multiple antibody binding sites, FMIA is less likely to lose performance when facing infections caused by various SARS-CoV-2 variants. The combination of antigens makes FMIA also less susceptible to possible cross-reactivity with antibodies produced against seasonal coronaviruses. Additionally, FMIA could be used to distinguish immune response induced by COVID-19 vaccination and recent SARS-CoV-2 infection.

The need for highly sensitive and specific antibody tests will continue in the future as new variants emerge fuelling further COVID-19 waves. Here we have described a reliable antibody assay for the simultaneous quantification of multiple SARS-CoV-2 antibodies, with excellent clinical performance and good comparability with NAb titers. We have also described the calibration of FMIA against a WHO international standard, which has been reported to reduce inter-laboratory variation notably [31]. However, we observed that calibrating FMIA and EIA only emphasised the differences between their results. In FMIA the results are calculated from a standard curve and in EIA from the ratio between a sample and two controls. Although standardisation of SARS-CoV-2 antibody assays is urgently needed, our results indicate only a limited value for the international standard in calibrating two assays whose test principles are quite different. The applicability and value brought by calibration should be considered when used in comparisons between methodologically diverse assays.

## Data Availability

The data that support the findings of this study are available from the corresponding author upon reasonable request.

## Authors’ contributions

CV, AH, AS, PJ and LK performed the laboratory analyses. AK, SM, LH and TD recruited the volunteers and managed serum sample collection. AS and CV developed and optimised the assay. AS, NE, CV and MM analysed the results. AS, JL, IJ and MM contextualised and AS, NE, AH, CV and MM drafted the manuscript. All authors approved the final version.

## Acknowledgements

Leena Saarinen, Marja Suorsa, Lotta Hagberg, Esa Ruokokoski, Tuomo Nieminen, Hanna Valtonen, Katja Lind, Heidi Hemmilä, Tiina Pulliainen, Marja-Liisa Ollonen, Minna Haanpää, Katri Keino, Riitta Santanen, Christel Pussinen, Mervi Nosa, Anne-Mari Pieniniemi and Anne Suominen.

