## Supplementary Material for "A highly sensitive and specific SARS-CoV-2 spike- and nucleoprotein-based fluorescent multiplex immunoassay (FMIA) to measure IgG, IgA and IgM class antibodies"

Supplementary data

### A detailed description of the SARS-CoV-2 FMIA

**Conjugation of microspheres.** SARS-CoV-2 RBD (product code: REC31849, the Native Antigen Company), SFL (product code: REC31868, the Native Antigen Company) and nucleoprotein (product code: REC31812, the Native Antigen Company) antigens were conjugated on the surfaces of MagPlex®-C superparamagnetic carboxylated microspheres (Luminex) by carbodiimide reaction as described previously [14].

**Preparation of plates.** Serum samples, the WHO reference serum and controls were diluted to PBS solution (pH 7.2) containing 1% (v/v) Tween20, 1% (w/v) bovine serum albumin, 0.5% polyvinyl alcohol and 0.8% (w/v) polyvinylpyrrolidone. 25 µl aliquots of diluted samples, reference serum and controls were added on black 96-well plates (Costar 3915) with 25 µl of PBS containing the three microsphere regions (1750 microspheres/region/well) each conjugated with a single type of SARS-CoV-2 antigen. The plates were incubated in RT with shaking at 600 rpm in the dark for 1 hour, after which unbound particles were washed away with a magnetic plate washer (ELx405 and 405TSRS, BioTek) by 2 x 200 µl of PBS containing 0.05% (v/v) Tween20. 50 µl 1:100 diluted IgG, IgA or IgM detection antibody conjugate was added to the wells. Detection antibodies used were R-Phycoerythrin-conjugated Affinipure Goat Anti-Human IgG, IgA or IgM Fcγ Fragment Specific (product code for IgG: 109-115-098, product code for IgA: 109-115-011, product code for IgM: 109-116-129, Jackson Immuno Research). The plates were incubated for 30 minutes in RT, in the dark with shaking at 600 rpm, after which unbound particles were washed away as described above. 80 µl of PBS was added to the wells and the plates were incubated for 5 minutes in RT, in the dark with shaking at 600 rpm before measurement of antibody levels with MAGPIX® system (Luminex). IgG, IgA and IgM antibody levels were measured from separate plates.

**Calculation of FMIA U/ml antibody concentrations.** IgG, IgA and IgM antibody levels were determined as antibody concentrations (FMIA U/ml) interpolated from 5-parameter logistic (5-PL) curves (xPONENT software version 4.2, Luminex) created from serially diluted (1:400–1:1638400) in-house reference sera each with two true duplicates. 1:400 dilution was given an arbitrary concentration of 100 FMIA U/ml. In addition to in-house reference serum and two blank wells, high and low concentration controls were included in each plate in true duplicates. The samples of the negative serum panel (n=402) were analysed as true duplicates diluted 1:100. The samples of the positive serum panel (n=147) were analysed as 1:100 and 1:1600 dilutions in true duplicates and the results were determined as the mean concentrations of two dilutions. Samples with antibody levels higher than the linear range of the reference curve were re-analysed with a higher serum dilution. The antibody concentration values under the limit of detection were given a value of one-half of the limit of detection (LOD).

### Evaluation of the analytical performance of FMIA.

**Precision.** The precision of FMIA was assessed by intra- and inter-assay variation as the percentage of the mean coefficient of variation (CV%) of antibody concentrations (FMIA U/ml). Intra-assay variation was determined as the mean CV% of all dilutions per antigen (n=70–91 replicates, 13 plates) and antibody class. Inter-assay variation was determined by comparing the levels of in-house reference serum analysed on the same day (IgM n=5, IgA n=6, IgG n=8) in different plates (IgM n=10, IgA n=12, IgG n=17). In addition, inter-assay variation was assessed by analysing the same five sera on five different days.

**Intermediate precision.** The reproducibility of the FMIA was assessed by comparing the results obtained by three laboratory technicians and as a variation caused by different batches of crucial reagents. All technicians analysed the same 19 samples included in the negative and positive serum panels. Mean CV% was calculated for all technicians together. Crucial reagents assessed were conjugated microspheres and detection antibodies. For IgA detection antibodies, two batches were used in the analysis of 21 samples and the mean CV% was calculated. For the detection of IgG antibodies, the sample sizes were 14 (N) and 33 (RBD and SFL). All IgM analyses were performed using the same batch of detection antibodies. The variation caused by different batches (n=4) of conjugated microspheres was assessed by a comparison of IgG antibody concentrations of two sets of microspheres at a time and then calculating the overall CV% per antigen between four batches. The number of samples differed between separate batch comparisons (n=13-31, 11-29 and 11-26 samples for N, RBD and SFL, respectively).

### Calibration against WHO International Standard.

The IgG specific in-house reference serum was calibrated against WHO International Standard (NIBSC code 20/136 [14]). The IgG specific concentrations against antigens N, RBD and SFL of the in-house reference serum were determined in an assay with the WHO international standard analysed in true duplicates with seven serial dilutions 1:100–1:1638400, with 1:100 being assigned with the concentration of 10 binding antibody units (BAU)/ml. The in-house reference serum was analysed as 8 true duplicates each serially diluted 1:100–1:1638400, and the analysis was repeated on two days. The mean concentration for each dilution of the in-house reference serum (n=22, 30 and 40 for nucleoprotein, RBD and SFL, respectively) was interpolated from the linear range (1:1600-1:409600) of the WHO international standard. In addition, these values determined for the in-house reference serum were used in an assay to measure the antibody concentrations of 30 serum samples (true duplicates, dilutions 1:100 and 1:1600), which were compared to the antibody concentrations obtained using the WHO international standard. The mean CV% of antibody concentrations of the sera (n = 28, 25 and 51 for nucleoprotein, RBD and SFL, respectively) was interpolated from the linear range of the WHO international standard, and the mean CV% of antibody concentrations below 20% and correlation R^2^≥0,95 was considered acceptable. Calibration factor was obtained for each antigen separately and used to convert FMIA U/ml results into BAU/ml.
