## Supplementary material for "A highly sensitive and specific SARS-CoV-2 spike- and nucleoprotein-based fluorescent multiplex immunoassay (FMIA) to measure IgG, IgA and IgM class antibodies": Table S1

Supplementary Table S1. Age and sex distributions of the participants that donated sera to pre-COVID-19 pandemic (negative serum panel, n=402), as part of the COVID-19 household study (positive serum panel, n=58) and as part of the Jalkanen et al. (2021) study [16] used in FMIA and EIA comparisons.

|  | Validation of CoV-2 FMIA | | | | FMIA and EIA comparisons | | | |
| --- | --- | --- | --- | --- | --- | --- | --- | --- |
|  | Negative serum panel | | Positive serum panel | | Vaccinated HCWs | | Patients | |
| Age group (years) | n (%) | % female | n (%) | % female | n (%) | % female | n (%) | % female |
| 1-10 | 100 (25) | 50 | 3 (5) | 0 | - | - | - | - |
| 11-20 | 38 (9) | 50 | 7 (12) | 57 | - | - | - | - |
| 21-30 | 36 (9) | 53 | 5 (9) | 20 | 2 (10) | 100 | 5 (25) | 40 |
| 31-40 | 38 (9) | 50 | 13 (22) | 62 | 6 (30) | 100 | 3 (15) | 67 |
| 41-50 | 38 (9) | 50 | 20 (34) | 65 | 9 (45) | 89 | 2 (10) | 50 |
| 51-60 | 38 (9) | 50 | 8 (14) | 63 | 2 (10) | 50 | 4 (20) | 25 |
| 61-70 | 38 (9) | 50 | 2 (3) | 50 | 1 (5) | 100 | 6 (30) | 50 |
| 71-80 | 38 (9) | 50 | - | - | - | - | - | - |
| 81-90 | 38 (9) | 50 | - | - | - | - | - | - |

HCWs = Healthcare workers
