## Supplementary material for "A highly sensitive and specific SARS-CoV-2 spike- and nucleoprotein-based fluorescent multiplex immunoassay (FMIA) to measure IgG, IgA and IgM class antibodies": Figure S1

Supplementary Figure S1. Distribution of positive serum panel’s days post-onset of symptoms at the time of sample collection.

| Days post-onset of symptoms | Number of samples |
| --- | --- |
| 0 | 0 |
|  | 5 |
| 20 | 23 |
|  | 26 |
| 40 | 33 |
|  | 10 |
| 60 | 1 |
|  | 0 |
| 80 | 0 |
|  | 1 |
| 100 | 6 |
|  | 8 |
| 120 | 21 |
|  | 8 |
| 140 | 3 |
|  | 2 |
