## Supplementary material for "A highly sensitive and specific SARS-CoV-2 spike- and nucleoprotein-based fluorescent multiplex immunoassay (FMIA) to measure IgG, IgA and IgM class antibodies": Table S2

Supplementary Table S2. Limits of quantification and detection of FMIA.

| Class | Antigen | LOQ | LOD |
| --- | --- | --- | --- |
| IgG | N | 0.0048* | 0.0094* |
|  | RBD | 0.0023* | 0.0057* |
|  | SFL | 0.0046* | 0.012* |
| IgA | N | 0.03** | 0.05** |
|  | RBD | 0.25** | 0.57** |
|  | SFL | 0.59** | 1.41** |
| IgM | N | 0.11** | 0.27** |
|  | RBD | 0.15** | 0.36** |
|  | SFL | 0.48** | 1.11** |

LOQ = limit of quantification, LOD = limit of detection,* = WHO-standard adjusted LOQ and LOD in BAU/ml, ** = non-adjusted LOD and LOQ in FMIA U/ml. N = SARS-CoV-2 (Wuhan-Hu-1) nucleoprotein, RBD = receptor binding domain of SARS-CoV-2 (Wuhan-Hu-1) spike glycoprotein, SFL = full length spike glycoprotein of SARS-CoV-2 (Wuhan-Hu-1).
