## Supplementary material for "A highly sensitive and specific SARS-CoV-2 spike- and nucleoprotein-based fluorescent multiplex immunoassay (FMIA) to measure IgG, IgA and IgM class antibodies": Table S3

Supplementary Table S3. Intra- and inter-assay variation of FMIA for each antibody class and antigen.

| Antibody class | Antigen | Intra-assay variation (CV%)^a^ | Inter-assay variation (CV%) | |
| --- | --- | --- | --- | --- |
|  |  |  | Within a day^b^ | Between days^c^ |
| IgG | N | 7 | 6 | 15 |
|  | RBD | 8 | 4 | 11 |
|  | SFL | 8 | 2 | 11 |
|  | Mean | 8 | 4 | 12 |
| IgA | N | 7 | 2 | 6 |
|  | RBD | 13 | 9 | 13 |
|  | SFL | 13 | 20 | 7 |
|  | Mean | 11 | 10 | 9 |
| IgM | N | 10 | 3 | 12 |
|  | RBD | 10 | 4 | 7 |
|  | SFL | 9 | 5 | 7 |
|  | Mean | 10 | 4 | 9 |

CV = coefficient of variation, N = SARS-CoV-2 (Wuhan-Hu-1) nucleoprotein, RBD = receptor binding domain of SARS-CoV-2 (Wuhan-Hu-1) spike glycoprotein, SFL = full length spike glycoprotein of SARS-CoV-2 (Wuhan-Hu-1), ^a^ = Calculated from antibody concentrations of in-house standards that were analysed in 13 assays as seven serial dilutions (1:400–1:1638400) each with two replicates. Results are the mean CV% of replicates within plates. ^b^ = In-house standard’s variation within one day. The mean of two replicates was used in the calculation of CV% between plates. Depending on the antibody class, values are based on 5–8 days and 10–17 assays (IgM: 5 days and 10 assays, IgA: 6 days and 12 assays, IgG: 8 days and 17 assays). c = Five sera analysed on five days as two replicates. The mean of two replicates was used in calculations of CV% between days.
