## Supplementary material for "A highly sensitive and specific SARS-CoV-2 spike- and nucleoprotein-based fluorescent multiplex immunoassay (FMIA) to measure IgG, IgA and IgM class antibodies": Table S4

Supplementary Table S4. Positive serum panel and the number and proportion of samples positive in MNT and FMIA.

|  | Number of samples | | Percentage of FMIA positive samples also positive in MNT  (number of all FMIA positive samples) | | |
| --- | --- | --- | --- | --- | --- |
| DPO | All | MNT positive | IgG positive* | IgA positive | IgM positive |
| <13 | 7 | 2 | 100% (3) | 100% (3) | 100% (3) |
| 13-20 | 21 | 20 | 100% (21) | 100% (21) | 100% (21) |
| 21-28 | 18 | 18 | 100% (18) | 100% (18) | 100% (18) |
| 29-36 | 30 | 30 | 100% (30) | 100% (30) | 90% (27) |
| 37-51 | 22 | 22 | 100% (22) | 95% (21) | 73% (16) |
| 52-150 | 49 | 45 | 100% (49) | 49% (24) | 38% (21) |
| Total | 147 | 137 | 100% (143) | 82% (117) | 73% (106) |

MNT = microneutralisation test, DPO = days post-onset of symptoms at sample collection.
* = Determined with spike glycoprotein antibody thresholds.
