## Supplementary material for "A highly sensitive and specific SARS-CoV-2 spike- and nucleoprotein-based fluorescent multiplex immunoassay (FMIA) to measure IgG, IgA and IgM class antibodies": Figure S2

Supplementary Figure S2. Spearman correlation (ρ) and significance (*p*) between IgA and IgM specific FMIA U/ml and EIA unit results. Dashed lines mark thresholds for positivity per antigen. S1 = SARS-CoV-2 spike glycoprotein S1 subunit, RBD = receptor binding domain of SARS-CoV-2 (Wuhan-Hu-1) spike glycoprotein. SFL = full-length spike glycoprotein of SARS-CoV-2 (Wuhan-Hu-1). One point may represent multiple samples (n=80).

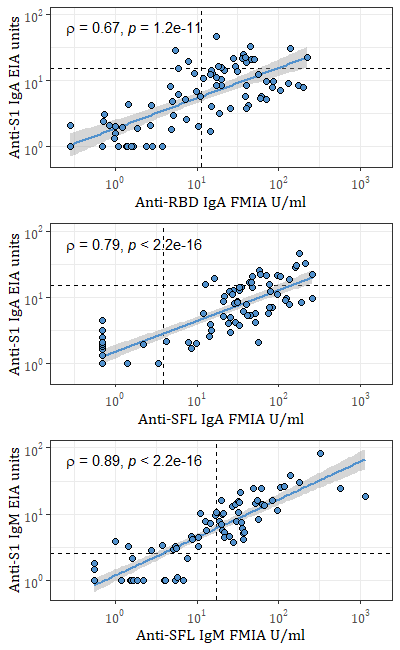
